# Enhancing Radiographic Diagnosis: CycleGAN-based methods for reducing cast shadow artifacts in wrist radiographs

**DOI:** 10.1101/2024.10.15.24315508

**Authors:** Stanley A Norris, Daniel Carrion, Michael Ditchfield, Manuel Gubser, Jarrel Seah, Mohamed K Badawy

## Abstract

**Objective:** We extend existing techniques by using generative adversarial network (GAN) models to reduce the appearance of cast shadows in radiographs across various age groups.

**Materials and Methods:** We retrospectively collected 12000 adult and pediatric wrist radiographs, evenly divided between those with and without casts. The test subset consisted of 100 radiographs with cast and 100 without cast. We extended the results from a previous study that employed CycleGAN by enhancing the model using a perceptual loss function and a self-attention layer.

**Results:** The CycleGAN model which incorporates a self-attention layer and perceptual loss function delivered the best quantitative performance. This model was applied to images from 20 cases where the original reports recommended CT scanning or repeat radiographs without the cast, which were then evaluated by radiologists for qualitative assessment. The results demonstrated that the generated images could improve radiologists’ diagnostic confidence, in some cases leading to more decisive reports. Where available, the reports from follow-up imaging were compared with those produced by radiologists reading AI-generated images. Every report, except two, provided identical diagnoses as those associated with follow-up imaging. The ability of radiologists to perform robust reporting with downsampled AI-enhanced images is clinically meaningful and warrants further investigation. Additionally, radiologists were unable to distinguish AI-enhanced from unenhanced images.

**Conclusion:** These findings suggest the cast suppression technique could be integrated as a tool to augment clinical workflows, with the potential benefits of reducing patient doses, improving operational efficiencies, reducing delays in diagnoses, and reducing the number of patient visits.

## 1. Introduction

Damaged limbs often require stabilisation using splints and casts made of resin, fibreglass or plaster (1). Removing the cast before imaging is impractical during follow-up radiographs monitoring the position or healing of fractures. The presence of a cast produces undesirable artefacts obscuring the visualisation of the bone structure and encompasses the examined anatomy. At the time of writing, there has been only a single study on cast suppression methods in extremity radiographs, by Hržić et al. (2). We attempt to reproduce these results on a larger data set and modify the model by adding a perceptual loss function and self-attention layer (3,4).

The field of medical imaging has significantly advanced due to the use and application of artificial intelligence (AI) methods in an extensive range of tasks. These models can be categorised as being vision, language, and vision-language models, depending on the nature of input and output data. In the context of cast suppression, the relevant vision model is known as a generative adversarial network (GAN). GANs have effectively been used for image processing tasks such as CT denoising, artefact reduction, radiotherapy planning, intermodality image synthesis, image reconstruction, data augmentation, image registration, classification, and inversion problems (5,6). When faced with an unpaired image problem, where there is no ground truth for the model to learn from, the cycle-consistent GAN (CycleGAN) model can be used. This approach can translate an image from a source domain to a target domain without paired images, so the distribution of generated images is indistinguishable from that of target images. A cycle consistency loss is introduced so the original image can be returned from the generated one. This method performs well for textural and colour changes in images but poorly for geometric changes, and the characteristics of the training data used limit its generality (7). Since the available casted radiographs do not have paired castless images, and a cast shadow mainly presents a textural change, the CycleGAN model is well suited to this problem.

One of the difficulties in quantitatively evaluating the model in the cast suppression problem is the lack of ground truth to which results can be compared. In some studies, a limited number of paired images were available and thus used for evaluation rather than training, such as for conversion of CT scans between reconstruction kernels (8), noise reduction in low-dose CT scans (9), or generation of CT images from CBCT images (10). For many different problems, CycleGAN models are evaluated by applying quantitative metrics such as Structural Similarity Index Metric (SSIM), histogram correlation, histogram intersection, Chi-squared distance, and Hellinger distance, which compare real and generated sets of images (2,11–14). The obvious limitation of these metrics is that spatial information is lost when converting an image to a histogram; for example, a circle and square of equal area and brightness would yield equivalent histograms despite representing distinct objects. In the original cast suppression study, a qualitative evaluation of the model was also undertaken by having radiologists of varying experience rank the generated images in terms of subjective quality. One study used a GAN to create lung nodules in CT images. It tested how well radiologists could distinguish real from fake and malignant from benign nodules (15). Another study used GANs to suppress bone in chest radiographs and tested for performance changes in radiologists’ ability to detect nodules (16). In what is perhaps the most compelling way of evaluating GAN models, the outputs are used as inputs to other AI models that have well-defined performance metrics. Examples include using GANs to improve segmentation and classification algorithms (17–24). A limitation in evaluating the CycleGAN method for cast suppression is a lack of precise application since there is no evidence that removing the cast shadow improves the quality or speed of extracting relevant clinical information in extremity radiographs. This presents an opportunity to bridge the gap between an exploratory study and an application that may yield tangible clinical benefits.

This study aims to extend the published CycleGAN method using an adult and pediatric dataset by incorporating a perceptual loss function and a self-attention layer. This study further evaluates the model’s performance through quantitative and qualitative analyses and investigates its potential for clinical application.

## 2. Materials and Methods

This study was exempt from Human Research Ethics Committee review as a retrospective quality improvement project. It was consistent with the NHMRC Ethical Considerations in Quality Assurance and Evaluation Activities (2014) guideline.

### 2.1 Image data acquisition

The images came from a large metropolitan hospital, including 30000 wrist radiographs taken between 2013 and 2023. We selected 10200 radiographs and divided them equally into groups of images with and without casts. First, original images were converted to 8-bit grayscale PNG files. Preprocessing included adding black pixels to images that weren’t square and rescaling to 512x512 pixels by Lanczos interpolation (25). The images were then grouped into training (n=10000) and testing (n=200) subsets. Half of the test images were without cast (n=100) and used as the reference for quantitative assessment, and the other half (n=100) were with cast and used to evaluate model performance.

### 2.2 Model architecture and implementation

As in the Hržić et al. study, the model is trained to optimise a loss function consisting of adversarial, cycle-consistency, and identity loss. The same training parameters are used for consistency as in previous publications (2,7) (cycle-consistency loss with weight λ =10, Adam optimiser with batch size 4, and learning rate α = 0.002, which was linearly reduced for the final 100 epochs). Due to computational limitations, and since the original study showed that the U-Net 512 architecture gave the best performance, we used this generator with 9 layers each for up- and downsampling. The details of the discriminators we applied are identical to those in the original study. The CycleGAN model was developed to include a perceptual loss (PL) function and self-attention layer (SAL). The PL function compares images based on differences between high-level image feature representations rather than differences between pixels, which may improve the quality of generated images (26). This study used a VGG16 network trained on the ImageNet dataset (27). The SAL added to the generator allows the network to consider the entire input when evaluating parts of the data, which may produce more coherent and higher-quality images by allowing the generator to capture intricate and global patterns in the images (28). The model was trained using Python (v3.10), Pytorch 2.4, and NVIDIA CUDA 12 libraries, on an NVIDIA Tesla P40 GPU with 24GB of VRAM. Details of training losses can be found in the supplementary information.

### 2.3 Quantitative image analysis

Applying the CycleGAN model to an image alters its associated pixel value histogram, particularly causing an increase in high-intensity values (2). In essence, the evaluation metrics quantify the similarity between two sets of histograms, which correspond to sets of histograms from real (H1) and generated (H2) castless images. In this study, we use histogram correlation (29), histogram intersection (30,31), Chi-squared distance (32), Hellinger distance (33), and Structural Similarity (SSIM) index (34,35) as quantitative metrics to assess the model. Details about quantitative metrics can be found in the supplementary information.

### 2.4 Qualitative image analysis

To probe the potential for clinical application, we first identified problematic images where the original radiology report suggested repeat imaging without the cast. We searched the Picture Archiving and Communication System (PACS) for 20 such cases. The model was then applied to these images to generate AI-enhanced versions. Three radiologists with 7, 10 and 39 years of experience were then asked to re-assess each case using the referral request or indication and the original and AI-enhanced images. The radiologists were blinded to the original reports, received all views associated with each imaging study, and could view the casted and decasted images side by side. The images were presented in a Google Form at low resolution (512×512), without the ability to adjust window and level settings. For each case, there was a field for the radiologists to write their report and a checkbox to indicate whether the AI-enhanced images improved their diagnostic confidence. A limitation of the qualitative assessment in the study by Hržić et al. is that observers ranked only generated images based on perceived quality, potentially turning the test into a “beauty contest” rather than focusing on diagnostic utility (36). To address this, the experiment compared the radiologists’ new reports against the original reports and categorised them into three outcomes:

1. The new report is essentially identical in identifying the presence or absence of fractures.
2. The new report is decisive about a fracture suspected in the original report.
3. The new report identifies a fracture not detected in the original report.

Where available, the new radiologist reports generated in this experiment were compared against reports from follow-up CT scans and radiographs without casts. Although the availability of such follow-up studies was limited, this comparison allowed us to establish a “gold standard” against which the new reports could be validated.

Each radiologist reviewed 60 image subsets via Google Forms as a separate Turing test. They were informed that the images may be all unenhanced, AI-enhanced, or a mixture of the two. Each image subset contains 15 real casted images, 15 generated castless images, 15 real castless images, and 15 generated casted images, all randomly selected. These images were unlabelled and randomly shuffled. The radiologists were allowed to zoom and pan within the image. They were also blinded to each other’s evaluation and were not shown any sample images before the assessment. The radiologists were asked to classify each image as unenhanced or AI-enhanced. This test determined whether radiologists can distinguish real from generated images, implying that the model can generate high-quality outputs if radiologists cannot detect the AI model (15). The generation of high-quality images could, for example, be leveraged as input for deep learning models.

### 2.5 Statistical analysis

First, a Shapiro-Wilk test was performed to determine that quantitative metrics were non-normally distributed. Since the histograms generated by each model are produced from the same set of real images, and the same set of reference images are used for comparison, we are dealing with paired data. Therefore, a Friedman test was performed since the data is non-normal, dependent, and consists of 4 groups. Post hoc analysis was done using the Wilcoxon signed-rank test, which must be applied for each combination of models with six possible combinations for four groups. In this case, a Bonferroni adjustment was applied to the threshold p-value for significance. A Chi-squared test for independence was performed to check if the distributions of report outcomes were different between radiologists. The Turing test assessments were tested for inter-rater reliability and inter-observer discrepancies via the Fleiss’ Kappa statistic and Chi-squared tests, respectively. The statistical analyses were performed using Python (v3.12.4).

## 3. Results

The dataset included 10,200 Radiographs, with an average patient age of 35 ± 28 years for those with casts and 35 ± 26 years for those without. Of these, 4,996 Radiographs were from male patients, with a mean age of 26 ± 21 years, and 5,204 Radiographs were from female patients, with a mean age of 44 ± 29 years.

The CycleGAN-PL-SAL model performed the best overall, with an SSIM of 0.5445 ± 0.1001, correlation of 0.9863 ± 0.0196, intersection of 191822 ± 28426, Chi-squared distance of 1136451 ± 2486185 and a Hellinger distance of 0.2902 ± 0.0867. The Friedman test yielded p-values of zero for all metrics. It thus implied that a Post Hoc test in the form of a Wilcoxon signed-rank test with a Bonferroni adjustment was needed to establish differences between the six comparisons between models. Most differences between models were statistically significant at the p = 0.05 level (Bonferroni adjusted to p < 0.009) in Wilcoxon signed-rank tests. The SSIMs between the CycleGAN and CycleGAN-PL (p = 0.07), CycleGAN and CycleGAN-PL-SAL (p = 0.11), and CycleGAN-PL and CycleGAN-PL-SAL (p = 0.03) were not significantly different. The histogram correlations between CycleGAN and CycleGAN-SAL (p = 0.01) and the Hellinger distances between CycleGAN-PL and CycleGAN-PL-SAL (p = 0.05) were not significantly different. The quantitative results are shown in Table 1 and Figure 1.

**Table 1:**
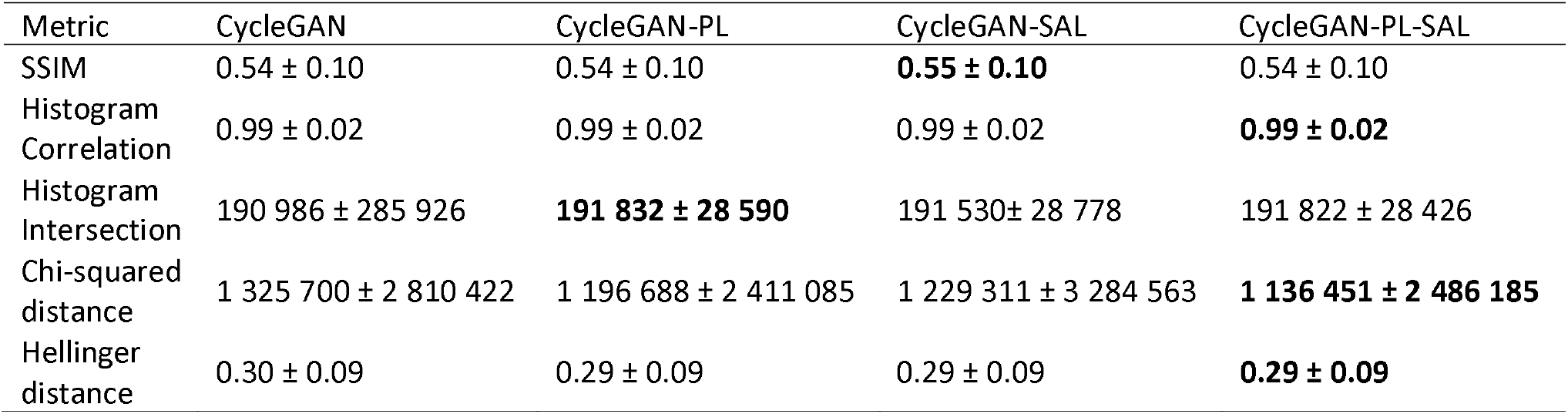
Quantitative comparison metrics with associated standard deviations for comparisons of the generated cast suppressed histograms against the reference castless test data set image histograms. The best results are given in bold.

| Metric | CycleGAN | CycleGAN-PL | CycleGAN-SAL | CycleGAN-PL-SAL |
| --- | --- | --- | --- | --- |
| SSIM | 0.54 ± 0.10 | 0.54 ± 0.10 | **0.55 ± 0.10** | 0.54 ± 0.10 |
| Histogram Correlation | 0.99 ± 0.02 | 0.99 ± 0.02 | 0.99 ± 0.02 | **0.99 ± 0.02** |
| Histogram Intersection | 190 986 ± 285 926 | **191 832 ± 28 590** | 191 530 ± 28 778 | 191 822 ± 28 426 |
| Chi-squared distance | 1 325 700 ± 2 810 422 | 1 196 688 ± 2 411 085 | 1 229 311 ± 3 284 563 | **1 136 451 ± 2 486 185** |
| Hellinger distance | 0.30 ± 0.09 | 0.29 ± 0.09 | 0.29 ± 0.09 | **0.29 ± 0.09** |

**Figure 1:**
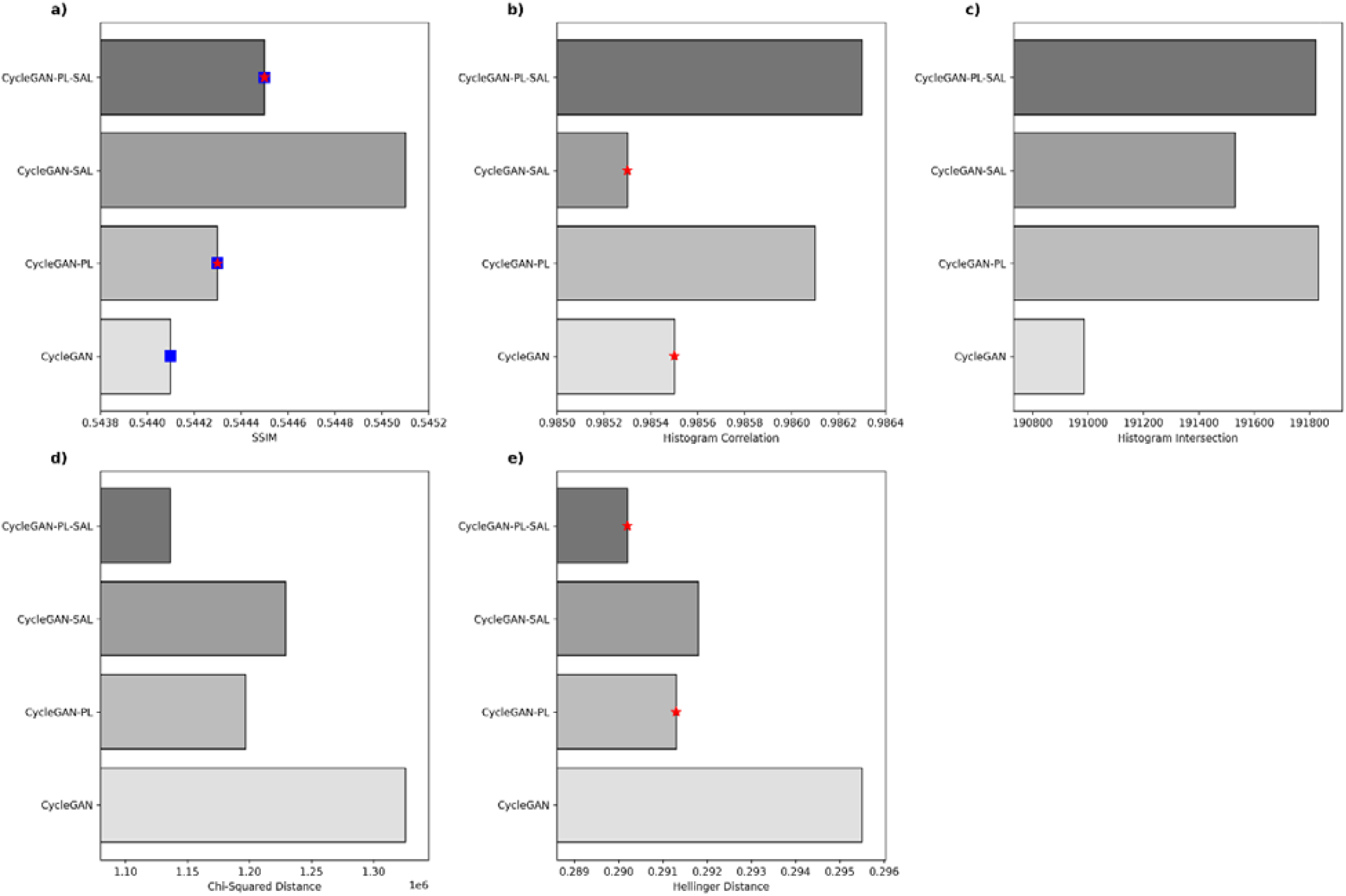
Bar charts displaying the quantitative metrics: a) SSIM, b) Histogram correlation and c) intersection, d) Chi-squared distance, and e) Hellinger distance. All differences between models were statistically significant at the Bonferroni-adjusted p < 0.009 level, apart from the 5 pairs indicated by the blue squares and red stars (p-values noted in main text).

As shown in Table 2, radiologists found that the AI-enhanced images improved diagnostic confidence in their reporting for 13, 16, and 11 of the 20 cases. In 4, 3, and 0 of the 20 cases, the radiologists reported more decisively and identified a fracture that was only suspected in the original report. In 2, 0, and 1 of the 20 cases, radiologists identified a fracture not mentioned in the original report. A summary of the fracture diagnoses in original reports are categorised and shown in Table 3. Since cases were selected because their original reports contained suggestions for repeat imaging, we performed a PACS search for repeat scanning. Follow-up imaging was available in 11 of the 20 cases, although one case with a follow-up radiograph had a report that still suggested further CT imaging. Of these 10 cases, 7 were followed up with X-rays and 3 were followed up with CT scans. Almost every report from radiologists reading AI-enhanced images had an identical diagnosis as that found in the report associated with follow-up radiographs and CT imaging. In a report that differed from follow-up imaging, one radiologist identified a “nondisplaced fracture of the proximal pole of the scaphoid bone”. Although all three radiologists correctly diagnosed the distal radius fracture confirmed by the follow-up CT scan, the follow-up report states that “the scaphoid is normal”. In the other report with a diagnosis that differed to follow-up imaging, one radiologist found that “linear lucency through the distal radius may represent a non displaced fracture although assessment is suboptimal”. In this case, the other two radiologists wrote “no fracture”, as determined in follow-up imaging. The Chi-squared test yielded a statistic of 6.47 (p=0.17), indicating that differences in the distribution of report outcomes were not statistically significant among radiologists.

**Table 2:** Summary results of the clinical application experiment, where diagnoses from each of 20 cases are placed into one of three categories. The far-right column indicates how many of these 20 images improve diagnostic confidence.

|  | Identical Report | More decisive report | New fracture identified | Improvement in diagnostic confidence |
| --- | --- | --- | --- | --- |
| Radiologist 1 | 14 (70%) | 4 (20%) | 2 (10%) | 13 (65%) |
| Radiologist 2 | 17 (85%) | 3 (15%) | 0 (0%) | 16 (85%) |
| Radiologist 3 | 19 (95%) | 0 (0%) | 1 (5%) | 11 (55%) |

**Table 3:**
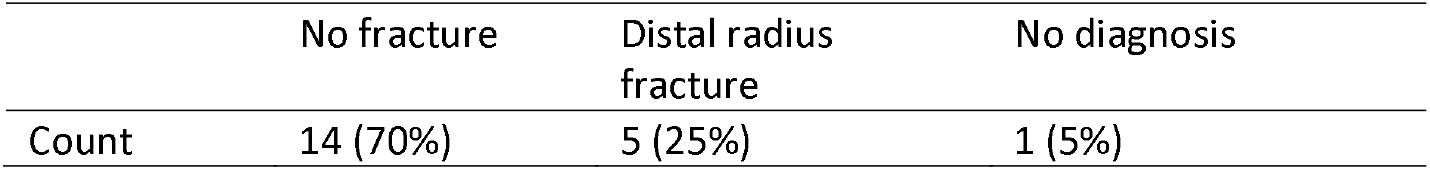
Summary of fracture diagnoses in the 20 cases identified by the PACS search for reports recommending follow-up imaging.

The Turing test experiment demonstrated that radiologists were typically unable to correctly identify AI-generated images. As shown in Figure 2, the radiologists classified 62% of the AI-enhanced images as unenhanced, while they correctly classified 87% of the unenhanced images. For this experiment, Fleiss’ Kappa statistic was 0.83 and a Chi-squared test yielded a statistic of 0.19 (p=0.91), showing a high level of agreement among raters, and that radiologists’ classifications were not significantly different from one another.

**Figure 2:**
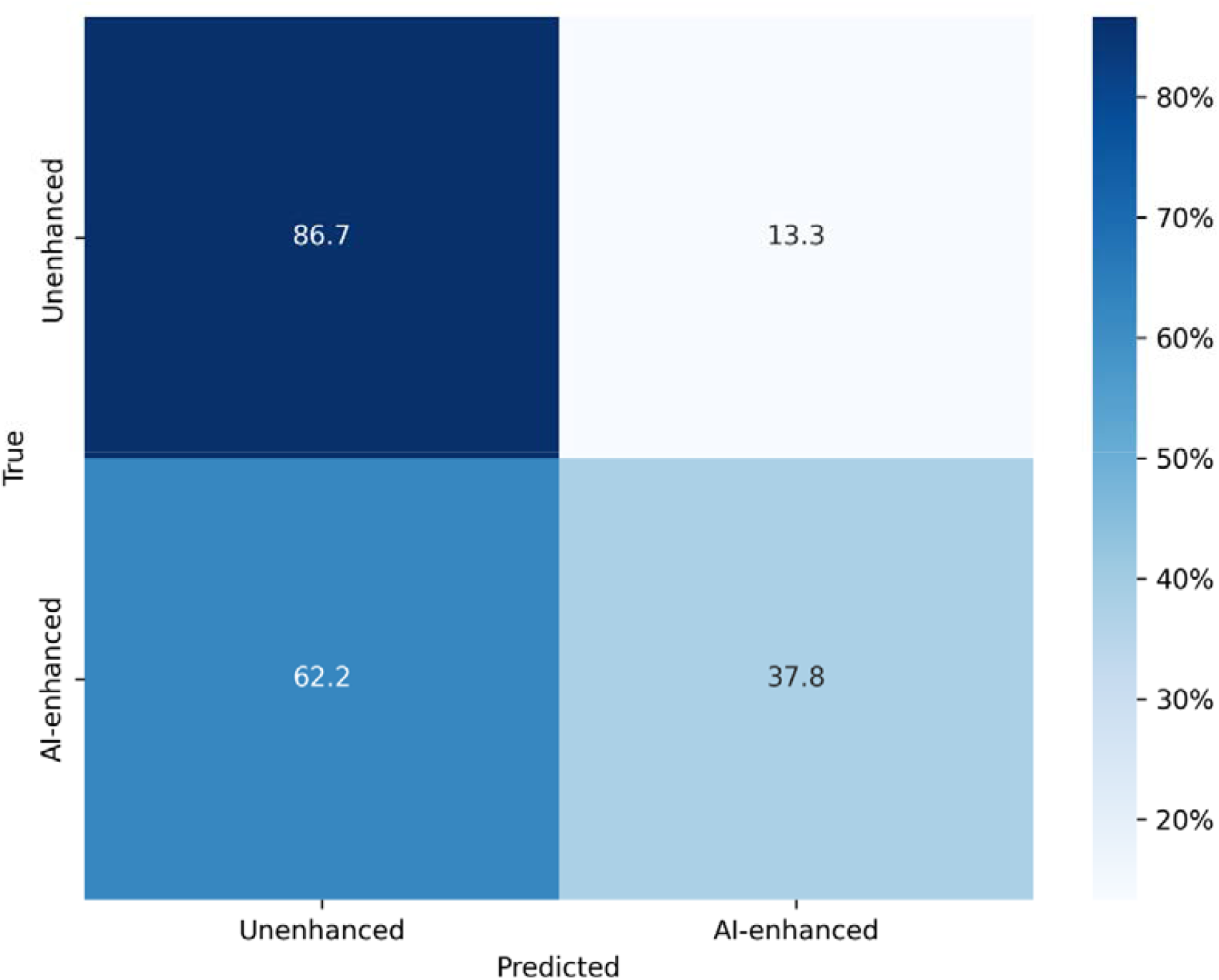
Confusion matrix of Turing Test results. The y-axis represents the nature of the displayed image and the x-axis represents the percentage of images radiologists classified into each category. Three radiologists evaluated 60 images each, giving a total of 180 observations. Radiologists’ assessments are grouped together since there was no significant difference between their classifications.

## 4. Discussion

In this study, we extended the method presented by Hržić et al. for suppressing cast artefacts in wrist radiographs by incorporating a perceptual loss function and a self-attention layer into the model. We investigated the effects of these additions on the model’s performance through quantitative and qualitative experiments. Additionally, our model was trained and evaluated on a dataset that required fewer preprocessing steps and was not limited to paediatric images. We quantitatively assessed the model outputs using standard image processing metrics and found that the enhanced model—including the perceptual loss function and self-attention layer—performed best overall. Although the CycleGAN-PL-SAL model showed relative improvement over the standard CycleGAN model, the quantitative metrics did not show absolute improvement compared to those reported by Hržić et al. However, these metrics are intrinsically non-reproducible because we used different sets of training and testing images. This inability to directly compare absolute values of quantitative metrics underscores a limitation in assessing models solely through these metrics.

The first qualitative test addressed a clinically relevant question and revealed that radiologists generally found that AI-enhanced images improved diagnostic confidence. In most cases, radiologists produced reports identical to the original ones, suggesting that the model does not generate significant artifacts or hallucinations. Notably, in some instances, radiologists provided decisive diagnoses of fractures that were only suspected in the original reports, and in three cases, they identified fractures that weren’t initially reported. This implies that the model may be suppressing casts effectively to uncover true underlying anatomy, demonstrating that thoroughly validated cast suppression could aid in diagnosis. One radiologist commented that the model sometimes made their task easier but never harder, expressing comfort in using the tool if it was well integrated with the PACS to reassure their diagnoses. This sentiment is reinforced by the finding that radiologists mostly produced reports with diagnoses identical to those from follow-up imaging. The one report that differed significantly to follow-up imaging may be partially attributed to the inherently greater diagnostic capability of CT imaging due to its tomographic spatial information. The other report that differed to follow-up imaging was inconclusive, indicating the AI-enhanced images were not sufficient to enable the radiologist to make a diagnosis. Given that all other reports provided diagnoses matching follow-up imaging, this suggests the model is not producing hallucinations but is generating reliable images for diagnosis. Furthermore, the demonstrated ability of radiologists to make robust diagnoses using downsampled images was particularly impressive.

The second subjective test in this study demonstrated that the model produces high-quality castless images, usually indistinguishable by radiologists from unenhanced images. Such realistic image generation could be leveraged for data augmentation, addressing data scarcity issues and class imbalances in datasets used to train other models. An objective validation approach could involve using the generated images as inputs for a well-established model to see if performance improves compared to using original images. For example, the model developed by Hembroff et al. can detect the presence of casts in wrist radiographs with high accuracy (37). This model could be tested with cast-suppressed images, objectively evaluating the quality of AI-generated images. These generated images could also serve as educational tools for radiology trainees and medical students.

This study is subject to various limitations relating to data, modeling, and performance evaluation methods, which present significant challenges for clinical implementation, particularly in establishing a robust validation method. While data availability is not an issue, it would not be difficult to collect a volume of data an order of magnitude larger. It is in fact the prohibitive computational costs that limit the number of images used in training. Training the model on data from multiple sources and institutions at native resolution would require significantly more computational resources. A major limitation of the clinical reporting assessment was using low-resolution images that could not be windowed or leveled, preventing the experiment from reflecting routine clinical conditions. Despite strong agreement between radiologists, some discrepancies in reporting were noted. The diagnostic reference standard we used demonstrated that the model can reliably generate images of sufficient diagnostic quality, which has implications for reducing cumulative population dose, improving operational efficiency, reducing delays to diagnoses, reducing patient visits, and improving radiologist workflows. However, this validation was limited in terms of scope. Despite these limitations, the model successfully generates high-quality cast-suppressed images. Examples of the model’s successes and failures are shown in Figure 3.

**Figure 3:**
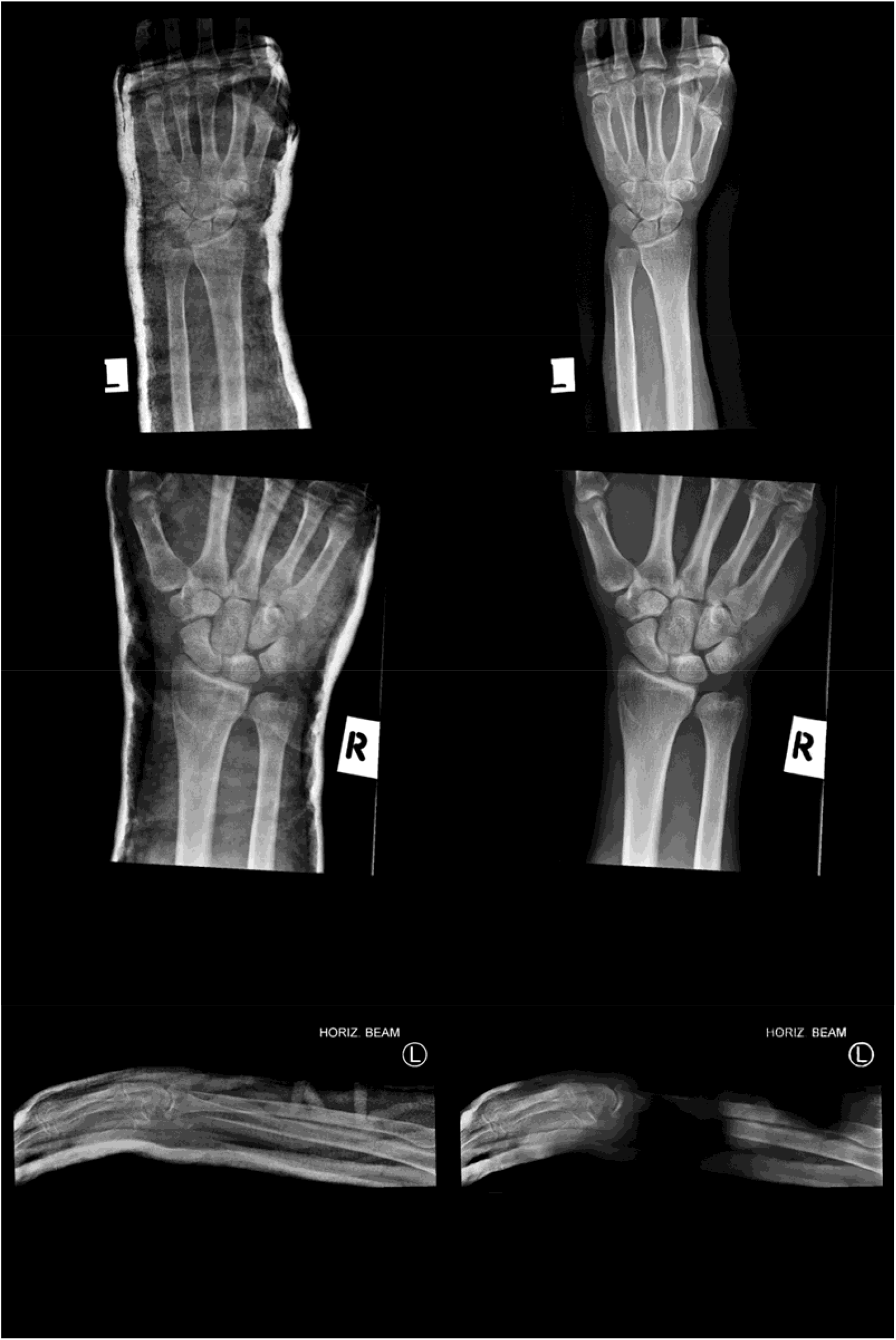
Collage showing several input images sampled from the test data set (left) and the associated outputs generated by the CycleGAN-PL-SAL model (right).

With access to greater resources, future work should focus on training the model on a greater volume of high-resolution images from various sources, and more comprehensively validating the results against follow-up imaging. The training and testing data should also include ankle radiographs, as an abundance of this data exists with and without casts. In-depth validation would also involve identifying a greater number of cases with follow-up imaging, and involving a larger cohort of radiologists in the assessment.

## 5. Conclusion

We extended a GAN-based method for suppressing cast artifacts in wrist trauma radiographs by incorporating a perceptual loss function and a self-attention layer into the model architecture. This enhancement led to relative improvements over the original method, as demonstrated by quantitative metrics on a large dataset that included adult and pediatric images. Qualitative assessments revealed that the model enhanced radiologists’ diagnostic confidence and that radiologists were typically unable to correctly identify AI-generated images. In some cases, the availability of both original and AI-generated images led radiologists to issue more decisive diagnoses for fractures that were previously only suspected, while also identifying new fractures that had not been mentioned in the original reports. The validation of a subset of reports against follow-up imaging demonstrates that the model can generate high-quality diagnostic images. Despite several limitations, this study lays the groundwork for developing a pipeline of AI models that can be built and eventually translated to the musculoskeletal radiology clinic. To the best of the authors’ knowledge, this is the first time that radiologists have reported on AI-generated, cast shadow suppressed wrist radiographs. This tool has demonstrated potential for reducing patient doses by avoiding repeat imaging, improving operational efficiency, reducing delays to diagnosis, reducing patient visits, and facilitating radiologist workflows.

## Data Availability

All data produced in the present study are available upon reasonable request to the authors

