## Supplementary information for "Enhancing Radiographic Diagnosis: CycleGAN-based methods for reducing cast shadow artifacts in wrist radiographs"

### **Quantitative metrics**

Applying the CycleGAN model to an image alters its associated pixel value histogram, particularly causing an increase in high-intensity values. In essence, the evaluation metrics quantify the similarity between two sets of histograms, which correspond to sets of histograms from real (H1) and generated (H2) castless images:

- Correlation, $\boldsymbol{d}_{\boldsymbol{\rho}}$, ranging from 1 to –1, the former relating to best histogram similarity:

$$d_{\rho}\left( H_{1},H_{2} \right) = \frac{\sum_{i} (H_{1}\left( i \right)-{H_{1}}^{-})(H_{2}\left( i \right)-{H_{2}}^{-})}{\sqrt{\sum_{i} {(H_{1}\left( i \right)-{H_{1}}^{-})}^{2}\sum_{i} {(H_{2}\left( i \right)-{H_{2}}^{-})}^{2}}}$$

where ${\boldsymbol{H}_{\boldsymbol{1}}}^{\boldsymbol{-}}$ and ${\boldsymbol{H}_{\boldsymbol{2}}}^{\boldsymbol{-}}$ are the average real and generated castless image histograms, respectively.

- Histogram intersection, $\boldsymbol{d}_{\boldsymbol{\cap}}$, which is a standard measure for histogram comparison where a larger intersection indicates greater similarity:

$$d_{\cap}\left( H_{1},H_{2} \right)= \sum_{i} min\left( H_{1}\left( i \right), H_{2}(i \right)).$$

- Chi-squared distance, $\boldsymbol{d}_{\boldsymbol{\chi}^{\boldsymbol{2}}}$, which represents the deviation of observed and expected frequencies:

$$d_{\chi^{2}}(H_{1},H_{2})= \sum_{i} \frac{{(H_{1}\left( i \right)- H_{2}\left( i \right))}^{2}}{H_{1}(i)}$$

where a smaller Chi-squared distance indicates more similar histograms.

- Hellinger distance, $\boldsymbol{d}_{\boldsymbol{H}}$, which quantifies the similarity of two distributions:

$$d_{H}\left( H_{1},H_{2} \right)= \sqrt{1- \frac{1}{\sqrt{H_{1}-H_{2}-N^{2}}}\sum_{i} \sqrt{H_{1}(i)\cdot H_{2}(i)}}$$

where N is the number of sampled images.

Finally, we also use the Structural Similarity (SSIM) index as a quantitative metric to assess the model. This metric is based on the assumption that humans are adapted to identify structural information from their visual perception. A higher SSIM value indicates signal structure retention between images and, thus, similarity.

### **Model training**

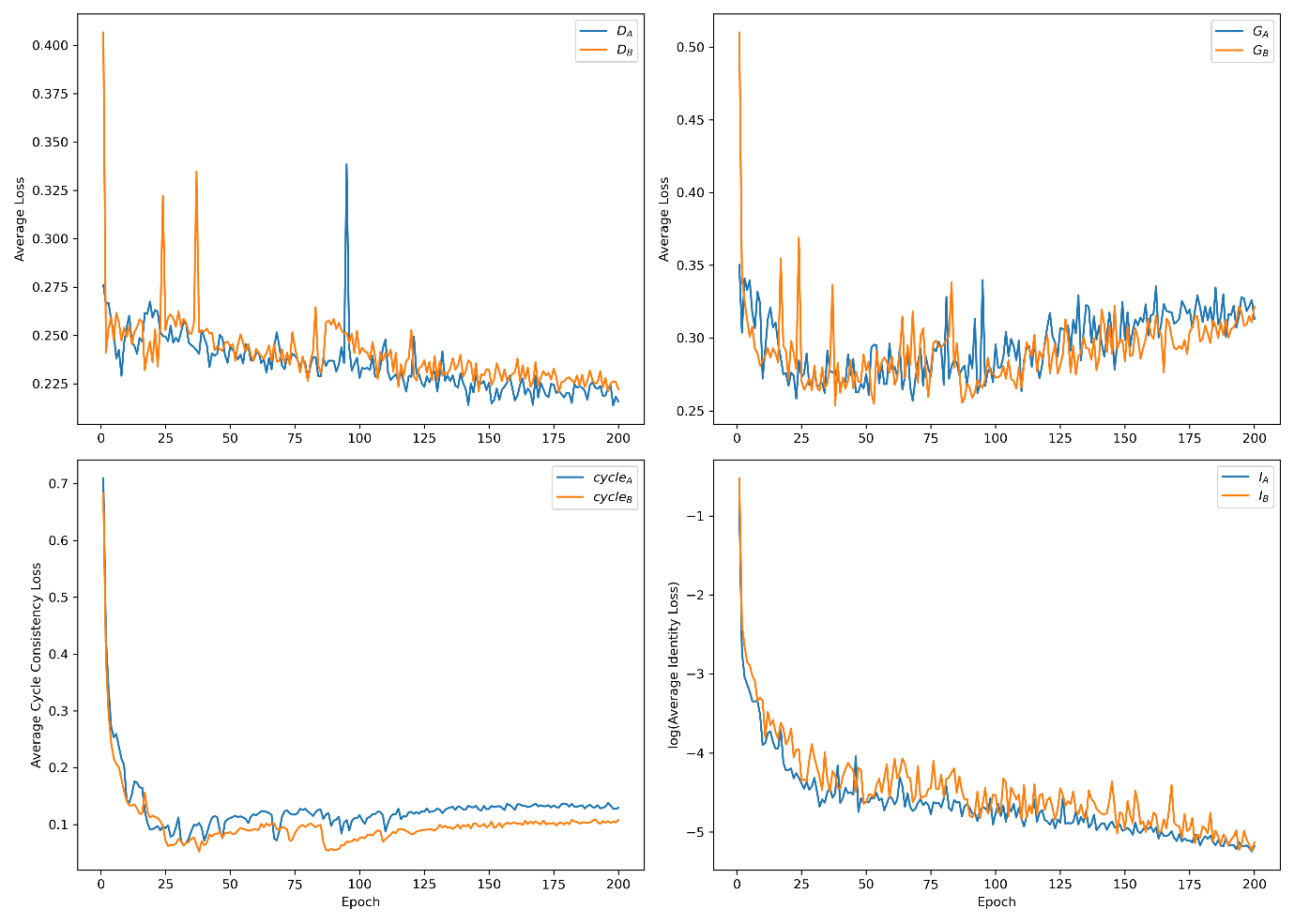

Figure 1: CycleGAN training losses over 200 epochs for Domains A (casts) and B (no cast). Top left: Discriminator losses. Top right: Generator losses. Bottom left: Cycle consistency losses. Bottom right: Identity losses decreases.

The top left graph shows the discriminator losses for both Domain A (D_A_) and Domain B (D_B_), which start high as the discriminators struggle to distinguish between real images and those generated by the CycleGAN. Over time, these losses decrease, indicating that the discriminators are learning to identify differences between real and generated images. The fluctuations are expected in GAN training due to the adversarial nature of the process.

In the top right graph, the generator losses for both domains (G_A_ and G_B_) decrease as the generators improve their ability to create realistic images. Some variability in the later epochs is typical of GAN models and indicates potential instability, though the overall trend suggests that the generated images are becoming more convincing. The slight increase at later epochs is not surprising for a GAN model as the generator is competing with the discriminator, which is effectively learning how to distinguish real and generated images.

The bottom left graph depicts the cycle consistency loss, measuring how well the model can translate an image from one domain to the other and back again. The decreasing and stabilizing losses for cycle_A_ and cycle_B_ show that the model is learning to preserve important information during domain translation.

The bottom right graph shows the natural logarithm of identity loss (I_A_ and I_B_), which ensures that images retain their key characteristics when passed through the generators within the same domain. A sharp decrease in the early epochs reflects the generators' rapid learning of this task, with further improvements seen as training progresses.

Overall, the graphs demonstrate that the CycleGAN is learning effectively, with strong improvements in discriminator, cycle consistency and identity losses. However, the increases in the generator losses suggest the model may still benefit from further fine-tuning to enhance training stability, which could be addressed in future work.
